# Are Longer Waits Associated with Lower Satisfaction and Worse Recovery? A Retrospective Observational Study of Hip and Knee Replacement in England

**DOI:** 10.64898/2026.10.28.26366729

**Authors:** Rohan Joseph, Shaun Danielli

**Author notes:** Correspondence to: Rohan Joseph.

## Abstract

**Objectives:** To examine whether longer waits for hip and knee replacement were associated with higher post-operative patient dissatisfaction and smaller patient-reported functional improvement, and whether worse post-operative satisfaction was associated with smaller functional improvement, in National Health Service (NHS) care in England.

**Design:** Retrospective observational study linking respondent-level Patient Reported Outcome Measures (PROMs) to provider-year Referral to Treatment (RTT) waiting times, with a supplementary comparison of the wait-dissatisfaction relationship in general practice.

**Setting:** NHS-funded hip and knee replacement in England, 2017, 2018, 2022 and 2023, excluding the pandemic-affected years 2019-2021.

**Participants:** 107,700 hip and 122,756 knee replacement PROMs respondents, and all 42 Integrated Care Boards (168 ICB-year observations) for the general practice comparison.

**Primary and Secondary Outcome Measures:** Post-operative dissatisfaction (fair or poor on a five-point satisfaction scale) and Oxford Score improvement (post-operative minus pre-operative score). Secondary outcomes were EQ-5D improvement and the full five-category satisfaction score.

**Results:** Each additional 30 days of average wait was associated with a 0.05 increase in the log-odds of post-operative dissatisfaction for hip (95% CI 0.03 to 0.08) and 0.04 for knee (0.03 to 0.06), and with smaller Oxford Score improvement (hip -0.26 points, -0.44 to -0.08; knee -0.11 points, -0.22 to -0.01). Mean Oxford Score improvement fell monotonically across satisfaction categories, from 26.11 points (hip) and 22.74 points (knee) among patients reporting excellent satisfaction to 2.62 and -0.01 points among those reporting poor satisfaction. In general practice, the wait-dissatisfaction association was close to zero but too imprecise to rule out an association of the size seen for hip and knee replacement.

**Conclusions:** In this focused surgical pathway, waiting times, post-operative satisfaction and patient-reported recovery were consistently associated, and satisfaction carried information about patient-reported recovery, not only about the experience of care. Performance frameworks that apply a common satisfaction measure across care settings should not assume it carries the same outcome content in each.

**Strengths and Limitations of this Study:**

- Respondent-level national PROMs data observed waiting time, post-operative satisfaction and a validated functional outcome (Oxford Score) for the same 107,700 hip and 122,756 knee replacement patients.
- Two wait exposures, average wait and share of pathways completed after 18 weeks, were each adjusted for comorbidities and fixed effects, with ordered logit and EQ-5D analyses as robustness checks.
- The wait exposure is a provider-year average across the whole trauma and orthopaedics treatment function, so it proxies rather than measures the procedure-specific wait.
- Case mix was adjusted only through 12 comorbidity flags; patient age and sex were not available in the PROMs extract.
- The design is observational, and because satisfaction is reported after recovery, the direction of the satisfaction-recovery association cannot be established.

## Introduction

Waiting times and patient dissatisfaction are widely used as measures of care quality, but their meaning is not always clear. Longer waits are usually interpreted as worse access, and dissatisfaction as worse patient experience. Both metrics are treated as core performance indicators across Organisation for Economic Co-operation and Development (OECD) countries.^1^ ^2^ However, whether they are associated with patient health, rather than only the process of care, has rarely been tested directly.

In this paper, we empirically examine the relationship between waiting time, patient dissatisfaction, and patient-reported functional improvement after hip and knee replacement in England. This setting allows all three measures to be observed for the same patients within a focused care episode. Using the Referral to Treatment (RTT) publications, which report the wait time from referral to admission, and the National Health Service (NHS) Patient Reported Outcome Measures (PROMs) publications, which report functional status pre- and post-procedure and post-operative dissatisfaction, we ask three questions. First, are longer waits associated with higher post-operative patient dissatisfaction? Second, are longer waits associated with smaller patient-reported functional improvement? Third, is worse post-operative satisfaction associated with smaller patient-reported functional improvement? As a supplementary comparison, we then ask whether the wait-dissatisfaction relationship extends to general practice, where the care episode is continuous rather than focused and patient satisfaction reflects communication and differences between patients as much as access.^3^ ^4^

The literature on these three questions is uneven. To begin, the association between waits and health gains is well established for orthopedic surgery. National UK PROMs data showed evidence of reduced health gains at longer waits, a meta-analysis of 89,996 patients extended this to a per-day deterioration in joint function and health-related quality of life before surgery, and a broader review found these associations particularly pronounced for orthopedic treatment.^5^ ^6^ ^7^ However, the evidence linking satisfaction to functional recovery is thinner and more mixed. In the National Joint Registry for England and Wales, pain and function were the dominant determinants of satisfaction after knee replacement.^8^ Satisfaction also reflected the fulfilment of pre-operative expectations after both procedures.^9^ ^10^ For hip replacement, satisfaction was associated with generic health-related quality of life improvement across 69,083 patients in the Swedish Hip Arthroplasty Register, whereas joint-specific scores could not predict satisfaction in a smaller sample in which most patients were satisfied.^11^ ^12^ The satisfaction measure itself is not standardised and has prompted calls for more consistent measurement, and international frameworks report satisfaction and functional outcomes as separate indicators without testing the association between them.^13^ ^14^ Finally, the association between waits and post-operative satisfaction has received the least attention. The only direct evidence comes from a single-center study of 192 knee replacement patients, in which waits over six months were associated with worse satisfaction and worse patient-reported outcomes.^15^

No previous study has jointly examined waiting times, patient satisfaction, and a validated measure of functional recovery for the same patients at national scale. We do so for hip and knee replacement in England, providing the first evidence beyond a single center on whether longer waits are associated with worse post-operative satisfaction. The joint test also shows whether two widely used performance metrics carry information about patient-reported health, rather than only about the process of care.

## Methods

### Study Design and Setting

We conducted a retrospective observational study of waiting times, patient dissatisfaction, and patient-reported functional improvement after hip and knee replacement in England, with a supplementary comparison of the wait-dissatisfaction relationship in general practice.

The main hip and knee analysis used respondent-level Patient Reported Outcome Measures (PROMs) observations linked to provider-year Referral to Treatment (RTT) waiting times, covering 2017, 2018, 2022, and 2023. We deliberately excluded the pandemic-affected years 2019 to 2021. Analysis year 2019 already captured the onset of the pandemic in both sources, because PROMs years are defined by financial year and General Practice Patient Survey (GPPS) fieldwork runs from approximately January to March of the following calendar year.

The linked sample contained 115,058 hip and 131,380 knee observations. All analyses used a single common analysis sample, which required non-missing values of the post-operative satisfaction response, the Oxford Score improvement, and the comorbidity controls, and excluded providers that could not be attributed to an Integrated Care Board (ICB), the regional body that plans and commissions NHS services. This gave 107,700 hip and 122,756 knee observations. The supplementary EQ-5D exhibits used the subset of this sample with non-missing EQ-5D improvement, comprising 99,037 hip and 112,792 knee observations. The provider-to-ICB attribution is described in Appendix E.

The supplementary general practice comparison used ICB by year observations from the GPPS and the Appointments in General Practice publications, covering all 42 ICBs in England in each analysis year, a total of 168 observations.

All analyses were descriptive and correlational. The paper follows the STROBE reporting guideline for observational studies.

### Data Sources

#### Referral to Treatment Waiting Times

Waiting time data were derived from the Referral to Treatment (RTT) publications, covering January 2017 to December 2023. The data reported monthly wait distributions for completed pathways across 104 weekly bins for each provider and treatment function. We subset to the trauma and orthopaedics treatment function (code C110), which is the treatment function under which hip and knee replacement is recorded, and which also covers other orthopaedic pathways.

Monthly bins were aggregated to provider-year cells. The average wait in days was calculated as the weighted mean of bin midpoints, using the share of completed pathways in each bin as weights, consistent with the published statistics’ own use of the binned distribution to estimate average waits.^16^ The highest bin is open-ended, and pathways in it were assigned a boundary value of 730 days, so the assigned value understates the true wait for the small share of pathways it contains. The share of pathways completed after 18 weeks was calculated as the sum of shares in the 18-week and higher bins.

#### Patient Reported Outcome Measures

Respondent-level data were obtained from the Patient Reported Outcome Measures (PROMs) publications for hip and knee replacement, covering financial years 2017/18 to 2023/24. The extract included each respondent’s pre- and post-operative Oxford Scores, pre- and post-operative EQ-5D indices, five-point post-operative satisfaction rating, and 12 comorbidity flags. Each respondent was linked to their provider’s RTT waiting times by provider code and PROMs year, defined as the first calendar year of the financial year. Providers were assigned a primary ICB using the provider-ICB crosswalk detailed in Appendix E.

#### General Practice Patient Survey

Patient dissatisfaction with general practice was obtained from the General Practice Patient Survey (GPPS) for survey years 2018, 2019, 2023, and 2024, which correspond to analysis years 2017, 2018, 2022, and 2023. The variable used was overall experience of the general practice, rated on a five-point scale, and dissatisfaction was defined as the combined share answering neither good nor poor, fairly poor, or very poor. Wording, scale, and dissatisfaction definition are consistent across the survey years used. The variable-code harmonisation is detailed in Appendix F.

#### Appointments in General Practice

General practice waiting time data were derived from the Appointments in General Practice publications, covering January 2017 to December 2023. We restricted to attended appointments delivered face to face, by telephone, or by video and online. Published waiting-time bands were converted to numeric midpoints in days, with the open-ended final band assigned 30 days, which is a lower bound on the true wait, and appointments with an unknown band were excluded. The ICB-year average wait was the appointment-weighted mean of these midpoints. Legacy Clinical Commissioning Group (CCG) level observations were mapped to current ICB geography using Office for National Statistics (ONS) Postcode Directory overlap weights, detailed in Appendix E.

### Measures

The first analysis outcome was post-operative patient dissatisfaction, coded as a binary variable equal to one for a satisfaction response of fair or poor on the five-point PROMs scale, and zero otherwise. The ordered five-category satisfaction score was used as a robustness outcome in Appendix A.

The second outcome was patient-reported functional improvement, represented by Oxford Score improvement in the main analysis and EQ-5D improvement as a supplementary check. Oxford Score improvement is the post-operative Oxford Hip or Knee Score minus the pre-operative score on a validated 0-48 scale, with higher values indicating greater functional recovery.^17^ ^18^ EQ-5D improvement is the post-operative EQ-5D index minus the pre-operative index, with higher values indicating greater improvement in generic health-related quality of life. Two wait exposures were used for hip and knee replacement. The first was the average wait, measured in 30-day units, and the second was the share of pathways completed after 18 weeks, measured in 10 percentage-point units. Both were constructed at the provider-year level.

For the supplementary general practice comparison, the outcome was the GPPS overall dissatisfaction share defined above and the exposure was the ICB-year average wait. The published appointment bands do not extend beyond 28 days, so no equivalent over-18-weeks measure can be constructed for general practice.

### Analytical Approach

The analysis proceeded in three parts. The first two each present a figure with a supporting regression estimated at the respondent level, reported to quantify the relationship shown in the figure. The third presents unadjusted box plots of Oxford Score improvement by satisfaction category.

First, we examined waiting times and patient dissatisfaction after hip and knee replacement. We presented binned scatter plots of residualised average wait against dissatisfaction, residualised on ICB fixed effects, year fixed effects, and respondent comorbidity controls, with supporting logit regressions clustered at the ICB level. Table A1 in Appendix A reports an ordered logit specification using the full five-category satisfaction outcome as a robustness check on the binary coding. The ordered logit did not converge with ICB fixed effects included, so that specification omits them and reports conventional standard errors.

Second, we examined waiting times and Oxford Score improvement, using the same construction with provider rather than ICB fixed effects, and ordinary least squares regressions with standard errors clustered at the provider level. Figure C1 in Appendix C reports the equivalent EQ-5D figure as a generic health-status check.

Third, we examined post-operative satisfaction and Oxford Score improvement, using box plots of Oxford Score improvement by satisfaction category with means marked. Figure D1 in Appendix D reports the equivalent EQ-5D figure.

For the supplementary comparison, hip and knee dissatisfaction were aggregated to the ICB-year level and the wait-dissatisfaction relationship was estimated for hip, knee, and general practice at the same unit of analysis, since general practice has no respondent-level linkage between waits and dissatisfaction. The specification was ordinary least squares weighted by the number of survey responses underlying each ICB-year dissatisfaction rate, with ICB and year fixed effects and standard errors clustered at the ICB level. Figure B1 and Table B1 in Appendix B report this comparison.

The regression specifications are:

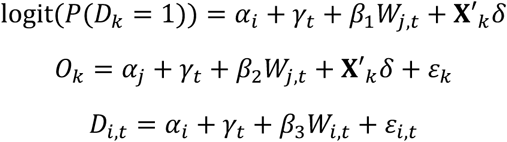

where *k* indexes respondent, *i* indexes ICB, *j* indexes provider, and *t* indexes year. *D_k_* is the binary dissatisfaction indicator for hip and knee, *O_k_* is Oxford Score improvement, *D_i_*_,*t*_ is the ICB-year dissatisfaction share in each setting (hip, knee or general practice), *W* is wait exposure, **X***_k_* is the vector of respondent comorbidity flags, and *α* and *γ* are unit and year fixed effects at the level indicated by their index.

### Patient and Public Involvement

Patients and members of the public were not directly involved in the design or conduct of this study. The analysis relied on publicly available administrative and secondary survey data. Patient-reported measures derived from PROMs were incorporated as key outcome indicators.

## Results

### Waiting Times and Patient Dissatisfaction

In Figure 1, we present binned scatter plots of residualised waiting time against residualised post-operative patient dissatisfaction for hip and knee replacement. Residualised dissatisfaction rose with residualised average wait across the wait distribution for both procedures. Longer waits were associated with higher post-operative patient dissatisfaction after hip and knee replacement.

**Figure 1.**
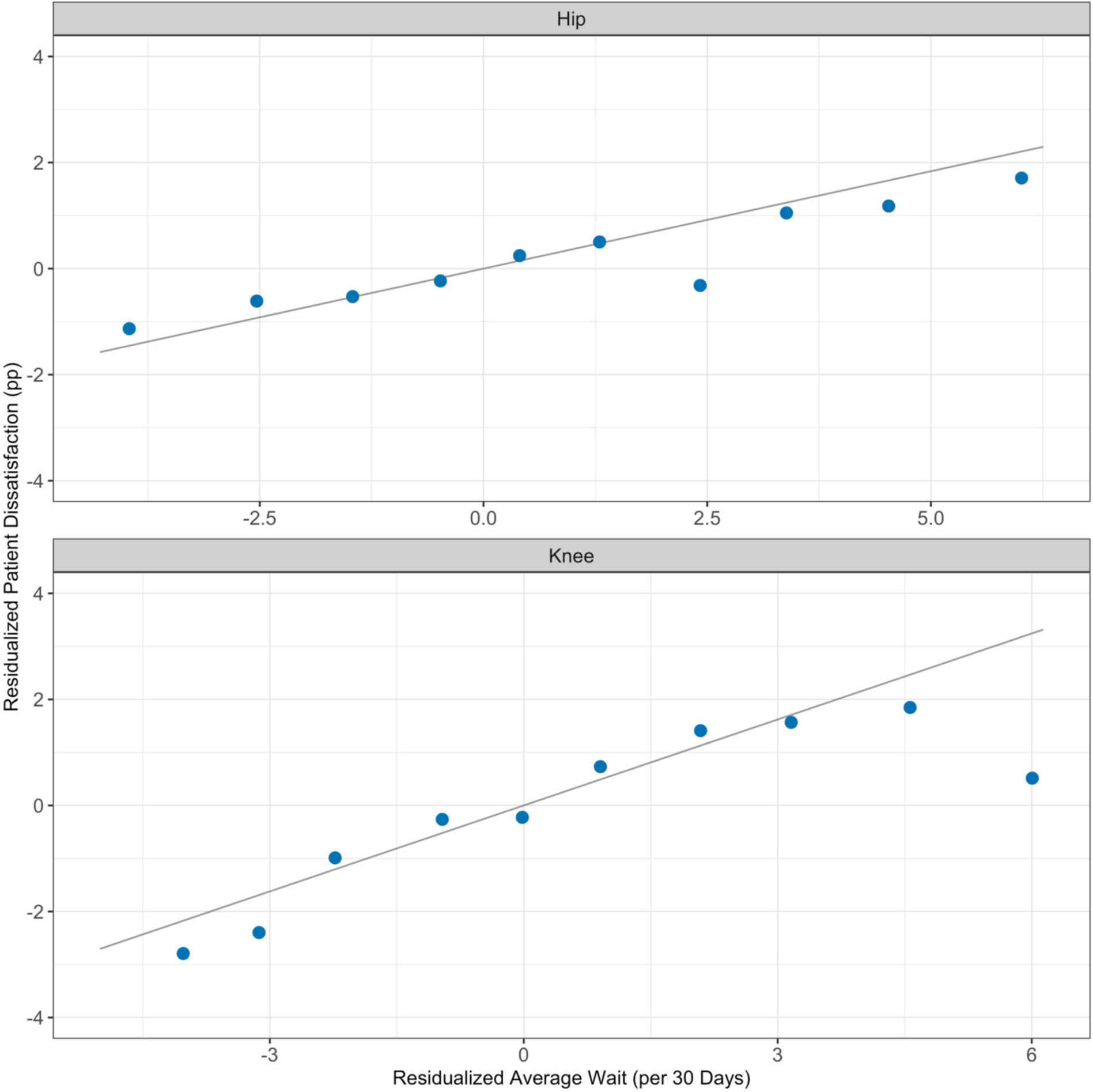
Average Wait and Patient Dissatisfaction after Hip Replacement and Knee Replacement. *Notes:* Binned scatter plots use PROMs respondent-level observations linked to provider-year RTT waits for 2017, 2018, 2022, and 2023, excluding 2019-2021, in the common analysis sample. Markers are bin means of residualised waiting time and post-operative dissatisfaction; bins with fewer than 20 observations are omitted. Variables are residualised on ICB and year fixed effects plus comorbidity controls. Waits are in 30day units and dissatisfaction is in percentage points.

Supporting logit regressions at the respondent level quantified the relationship (Table 1). Each additional 30 days of average wait was associated with a 0.05 increase in the log-odds of post-operative dissatisfaction for hip (95% CI: 0.03, 0.08) and 0.04 for knee (95% CI: 0.03, 0.06). Each additional 10 percentage points in the share of pathways completed after 18 weeks was associated with a 0.07 increase in the log-odds for hip (95% CI: 0.04, 0.11) and 0.05 for knee (95% CI: 0.03, 0.07). All four estimates were positive and their intervals excluded zero. Table A1 in Appendix A reports the ordered logit specification using the full five-category satisfaction outcome and shows the same graded pattern.

**Table 1.**
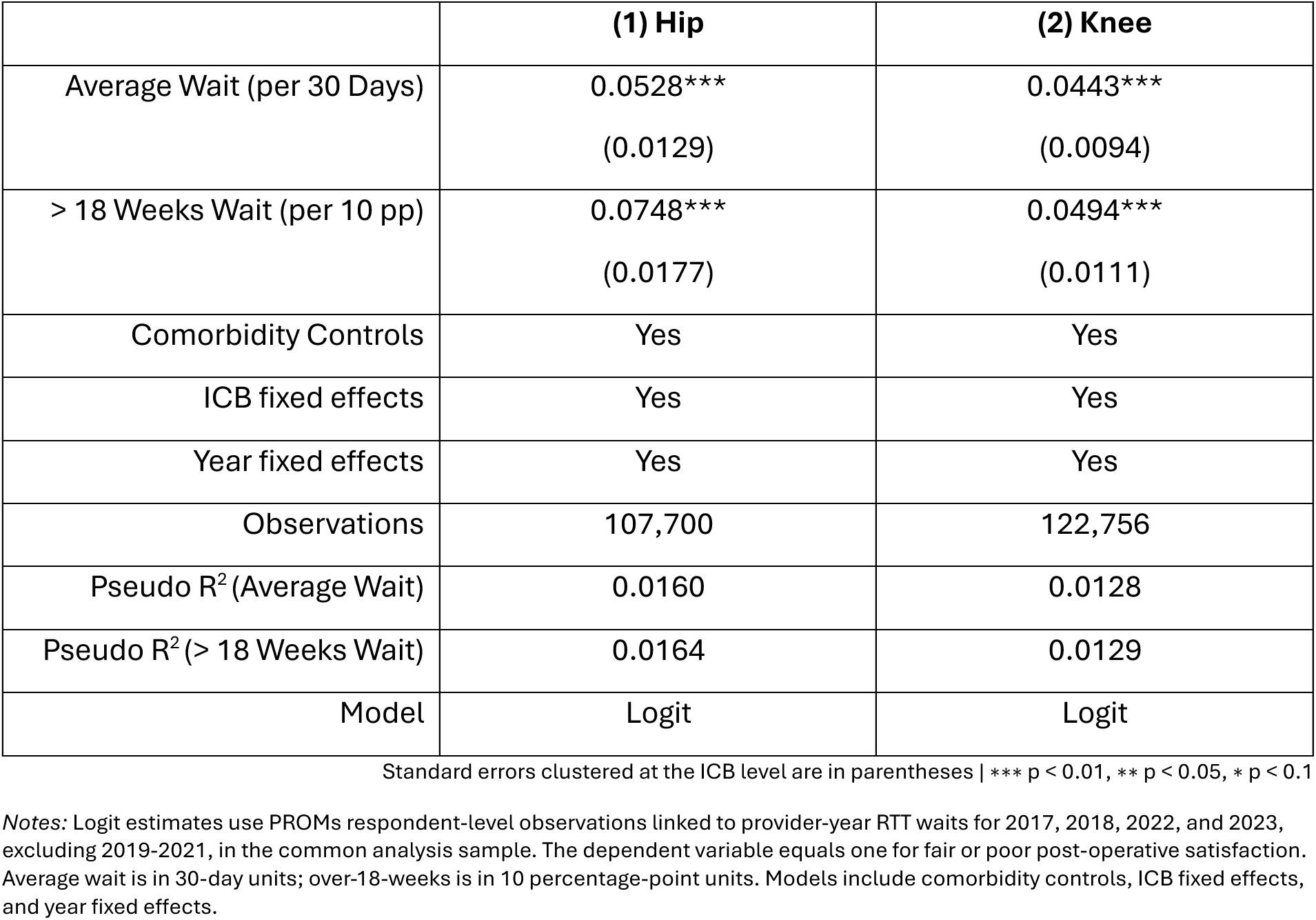
Logit Regression Estimates of the Association between Waiting Times and Post-Operative Patient Dissatisfaction after Hip and Knee Replacement.

### Waiting Times and Oxford Score Improvement

In Figure 2, we present binned scatter plots of residualised waiting time against residualised Oxford Score improvement for hip and knee replacement. Residualised Oxford Score improvement fell with residualised average wait across the wait distribution for both procedures. Longer waits were associated with smaller patient-reported functional improvements after hip and knee replacement. Figure C1 in Appendix C shows the equivalent pattern for EQ-5D improvement.

**Figure 2.**
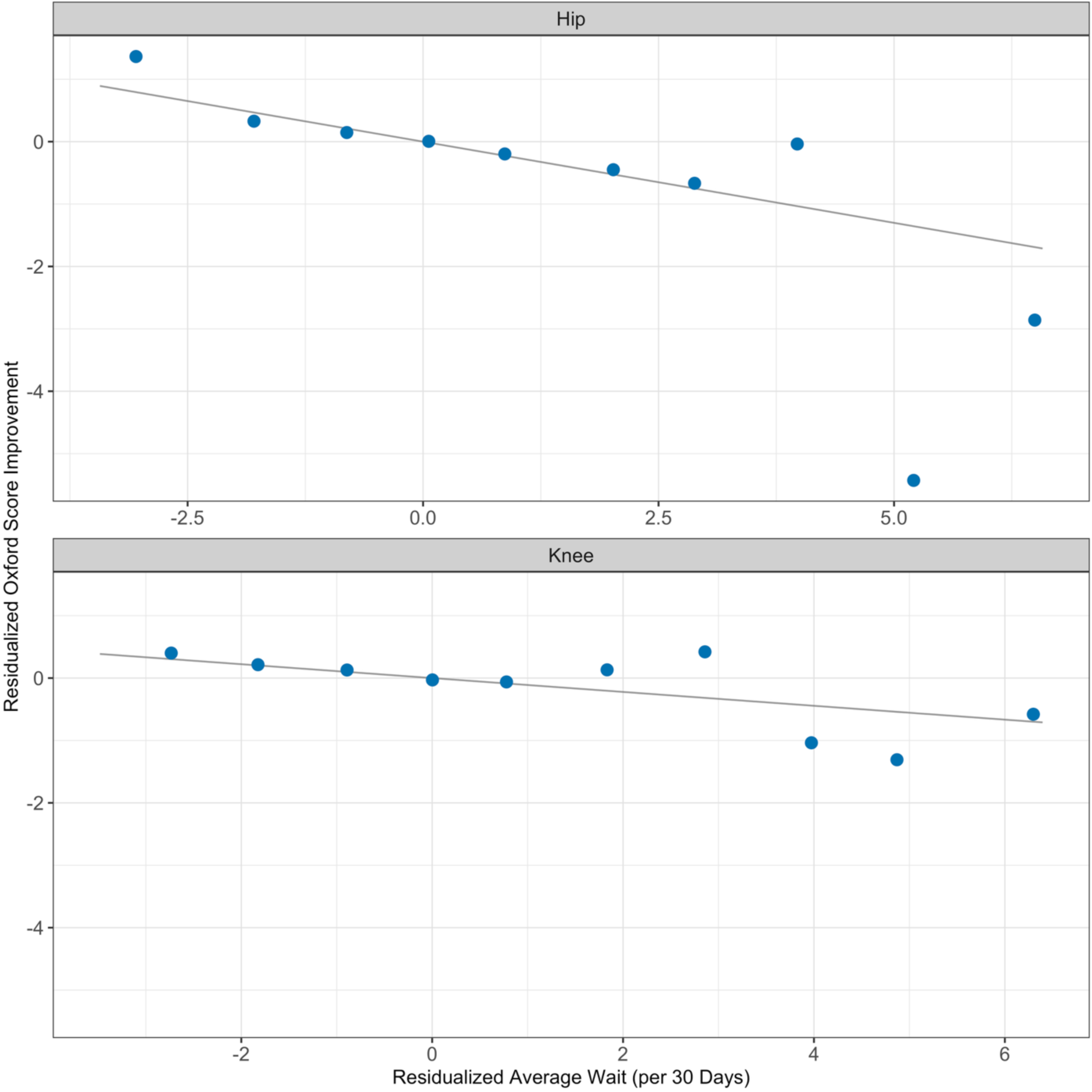
Average Wait and Oxford Score Improvement after Hip and Knee Replacement. *Notes:* Binned scatter plots use PROMs respondent-level observations linked to provider-year RTT waits for 2017, 2018, 2022, and 2023, excluding 2019-2021, in the common analysis sample. Markers are bin means of residualised waiting time and Oxford Score improvement; bins with fewer than 20 observations are omitted. Variables are residualised on provider and year fixed effects plus comorbidity controls. Oxford Score improvement is post-operative minus pre-operative score; waits are in 30-day units.

Supporting ordinary least squares regressions at the respondent level quantified the relationship (Table 2). Each additional 30 days of average wait was associated with a -0.26-point change in Oxford Score improvement for hip (95% CI: -0.44, -0.08) and -0.11 for knee (95% CI: -0.22, -0.01). Each additional 10 percentage points in the share of pathways completed after 18 weeks was associated with a -0.36-point change for hip (95% CI: -0.57, -0.16) and -0.18 for knee (95% CI: -0.30, -0.05). All four associations indicated smaller improvements at longer waits and their intervals excluded zero.

**Table 2.** OLS Estimates of the Association between Waiting Times and Oxford Score Improvement after Hip and Knee Replacement.

|  | (1) Hip | (2) Knee |
| --- | --- | --- |
| Average Wait (per 30 Days) | -0.2602***<br>(0.0898) | -0.1110**<br>(0.0535) |
| > 18 Weeks Wait (per 10 pp) | -0.3639***<br>(0.1057) | -0.1751***<br>(0.0616) |
| Comorbidity Controls | Yes | Yes |
| Provider fixed effects | Yes | Yes |
| Year fixed effects | Yes | Yes |
| Observations | 107,700 | 122,756 |
| R<sup>2</sup> (Average Wait) | 0.0204 | 0.0206 |
| R<sup>2</sup> (> 18 Weeks Wait) | 0.0206 | 0.0207 |
| Model | OLS | OLS |

Standard errors clustered at the provider level are in parentheses | \*\*\* p < 0.01, \*\* p < 0.05, \* p < 0.1
Notes: OLS estimates use PROMs respondent-level observations linked to provider-year RTT waits for 2017, 2018, 2022, and 2023, excluding 2019-2021. The dependent variable is Oxford Score improvement, defined as post-operative minus pre-operative score. Average wait is in 30-day units; over-18-weeks is in 10 percentage-point units. Models include comorbidity controls, provider fixed effects, and year fixed effects.

### Patient Satisfaction and Oxford Score Improvement

In Figure 3, we present box plots of Oxford Score improvement by post-operative satisfaction category, with means marked. Worse post-operative satisfaction was associated with smaller Oxford Score improvements. For hip replacement, mean Oxford Score improvement was 26.11 points among respondents reporting excellent satisfaction (95% CI: 26.04, 26.19) and 2.62 points among those reporting poor satisfaction (95% CI: 2.15, 3.08). The corresponding means for knee replacement were 22.74 points (95% CI: 22.65, 22.82) and -0.01 points (95% CI: -0.26, 0.23). The gradient was monotonic across all five categories in both procedures. Figure D1 in Appendix D shows the equivalent pattern for EQ-5D improvement.

**Figure 3.**
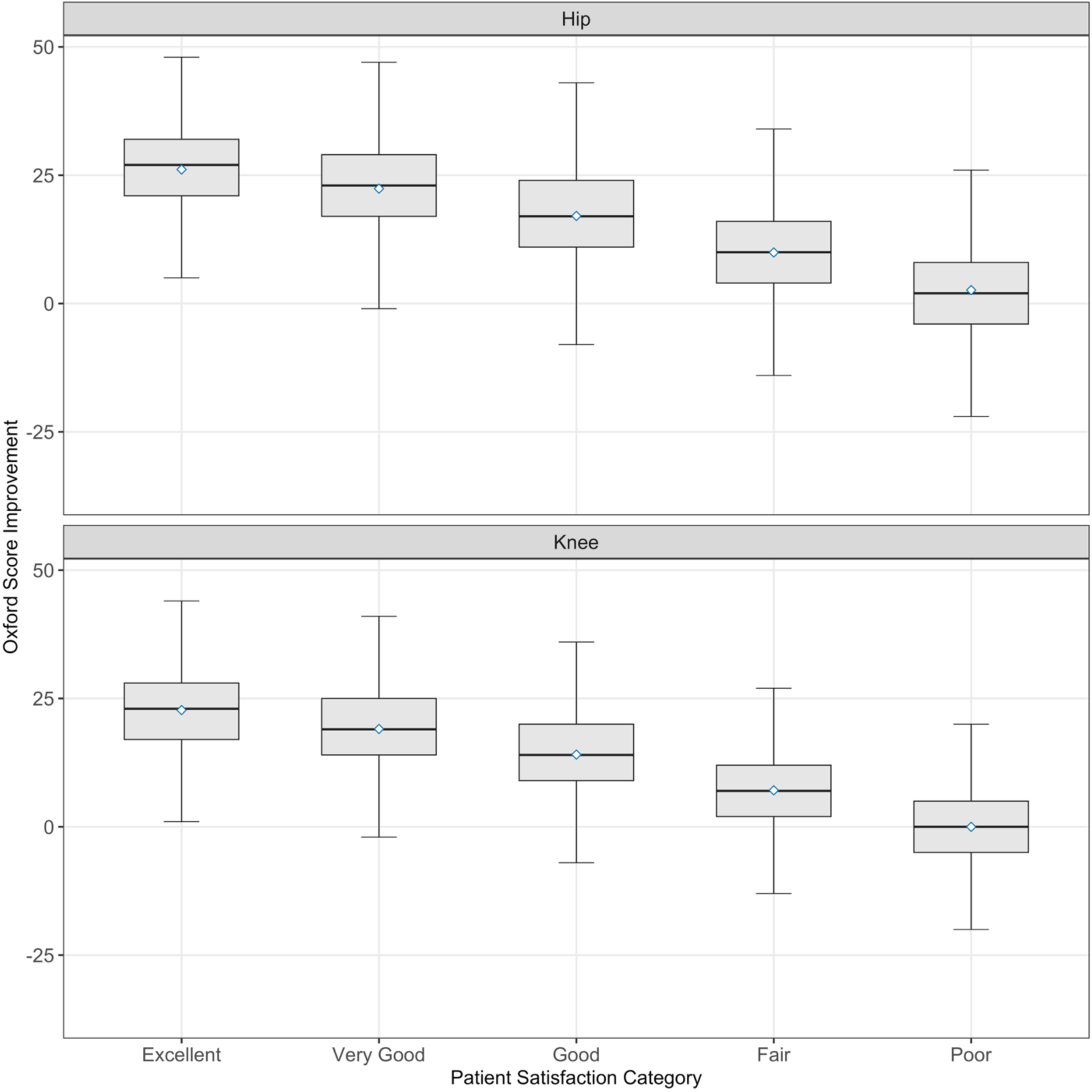
Oxford Score Improvement by Post-Operative Patient Satisfaction Category after Hip and Knee Replacement. *Notes:* Box plots use PROMs respondent-level observations linked to provider-year RTT waits for 2017, 2018, 2022, and 2023, excluding 2019-2021, in the common analysis sample. Oxford Score improvement is post-operative minus pre-operative score. Boxes show the interquartile range, whiskers show adjacent values, and diamonds show means.

### Supplementary Comparison with General Practice

We then compared the wait-dissatisfaction relationship in hip and knee replacement with the equivalent relationship in general practice. Hip and knee dissatisfaction were aggregated to the ICB-year level, and all three settings were estimated on a common footing, giving 168 observations per setting (Appendix B, Table B1). Aggregation to this level reduced the precision available to detect within-ICB variation.

In this specification, the association between average wait and patient dissatisfaction was 0.0048 percentage points per additional day for hip (95% CI: -0.0045, 0.0141), 0.0131 for knee (95% CI: 0.0037, 0.0225), and -0.0117 for general practice (95% CI: -1.4171, 1.3936). The knee estimate remained positive and its interval excluded zero. The hip estimate retained the expected positive sign, but its interval included zero. The general practice estimate was close to zero and imprecisely estimated, with an interval wide enough to contain both a null and an association of the magnitude estimated for hip and knee replacement. Figure B1 in Appendix B shows the general practice relationship, which was flat across the wait distribution.

## Discussion

Longer waits after referral for hip and knee replacement were associated with higher post-operative patient dissatisfaction and with smaller Oxford Score improvements. In addition, worse post-operative satisfaction was associated with smaller Oxford Score improvements, with mean improvement falling monotonically from excellent to poor across both procedures. Taken together, post-operative satisfaction after hip and knee replacement carried information about patient-reported recovery, not only about the experience of care.

The association between longer waits and smaller functional gains is consistent with the established evidence.^5^ ^6^ ^7^ The association between longer waits and higher post-operative dissatisfaction was consistent with the single-center knee replacement evidence, observed here at national scale and for both procedures.^15^ Our estimates also separate hip from knee, and distinguish the average wait from the share of pathways completed after 18 weeks, which places the association in the tail of the wait distribution as well as at its mean. The magnitude is modest. The minimal important change in the Oxford Hip and Knee Scores has been estimated at approximately 8 and 7 points respectively for an individual patient.^19^ The associations of 0.2602 points per 30 days for hip and 0.1110 points for knee may therefore be better read as a shift in the distribution of recovery across a large patient population than as a change an individual patient would notice.

The satisfaction-Oxford association reflects convergence between two different measures rather than one measuring the other. Satisfaction is a global judgement of the procedure, reported after recovery, and reflected the fulfilment of pre-operative expectations as well as recovery itself.^9^ ^10^ The Oxford Score is a validated joint-specific assessment of pain and function. That the two were associated with each other across the full five-category range, for the same respondents, was consistent with the pain-and-function evidence from the National Joint Registry and with the Swedish registry association between satisfaction and generic health improvement.^8^ ^11^ The estimates also contrasted with the null result from the smaller sample in which most patients were satisfied.^12^ These results add a national, joint-specific observation for both procedures to that mixed evidence. They also speak to the concern that satisfaction measurement in this setting is not standardised, since whatever the PROMs satisfaction question captures, it was associated with a validated functional outcome.^13^

The main implication concerns how patient-reported measures are used to assess performance. In a focused care episode in which the wait is a dominant feature, a single satisfaction item was consistently associated with a validated measure of recovery across the full range of the satisfaction scale. In this setting, patient satisfaction carried information about patient-reported health, and not only about the process of care. Where international frameworks report satisfaction and functional outcomes as separate indicators, the two are more closely related in a focused pathway than that separation suggests.^14^

The supplementary comparison with general practice sets a boundary on that reading, though a weak one. Aggregated to the ICB-year level the general practice association was close to zero, but the interval was wide enough to contain both a null and an association of the magnitude estimated for hip and knee replacement. So, the comparison is consistent with the relationship being specific to narrow care episodes without establishing it. What is clearer is what the two measures summarise. Post-operative PROMs satisfaction refers to a single procedure. General practice dissatisfaction pools access, continuity, communication, appointment type, expectations, case mix, and the ongoing relationship with the practice, and communication and differences between patients have been found to dominate responses in that setting.^3^ ^4^ Satisfaction measured over a broad and continuing care relationship should not be assumed to carry the same outcome content as satisfaction measured over one procedure.

Several limitations apply. All analyses are associational and no causal claims can be made, and because post-operative satisfaction is reported after recovery the satisfaction-Oxford relationship cannot separate directions. The wait exposure is the provider-year average across the trauma and orthopaedics treatment function, which covers pathways beyond hip and knee replacement, so it proxies the procedure-specific wait rather than measuring it and waits above 104 weeks were assigned a 730-day boundary value. Case mix was adjusted through 12 comorbidity flags, and the PROMs extract used did not include patient age or sex. The dissatisfaction and Oxford Score specifications use ICB and provider fixed effects respectively, so they do not hold the same factors fixed and their magnitudes are not directly comparable. Providers were assigned to a single primary ICB rather than proportionally allocated, and the assignment was built from RTT activity in the current-ICB era of the publication, which begins after the 2017 and 2018 analysis years. The general practice comparison is estimated at the ICB-year level with a compressed within-ICB wait range, and its imprecision limits what the cross-setting comparison can support. Finally, the pandemic-affected years were excluded, so the results speak to the 2017-2018 and 2022-2023 windows. Each of these limitations affects how far the reported associations should be pushed.

## Conclusion

In NHS-funded hip and knee replacement in England between 2017 and 2023, excluding the pandemic-affected years, longer waits were associated with higher post-operative dissatisfaction and smaller Oxford Score improvements, and worse satisfaction was associated with smaller improvements. Across this focused surgical pathway, waiting times, patient satisfaction, and patient-reported recovery were consistently associated, and post-operative satisfaction was associated with patient-reported recovery as well as the experience of care. No equivalent association was found between waits and dissatisfaction in general practice, although that estimate was imprecise. Performance frameworks that apply a common satisfaction measure across settings should therefore account for the breadth of the care episode, and future research should test whether these relationships hold in other focused clinical pathways.

## Statements

### Contributors

RJ and SD conceived the study. RJ led the data analysis and first draft of the manuscript. All authors contributed to subsequent drafting of the manuscript and accept full responsibility for, and have read and approved, the final manuscript. RJ is the guarantor. The corresponding author attests that all listed authors meet authorship criteria and that no others meeting the criteria have been omitted. The views expressed in this manuscript are those of the authors and not necessarily those of their institutions.

### Patient and Public Involvement

Patients and/or the public were not involved in the design, or conduct, or reporting, or dissemination plans of this research. Refer to the Methods section for further details.

### Patient Consent for Publication

Not applicable.

### Ethics Approval

This study used publicly available administrative data and record-level patient-reported data containing no identifiable patient information. Ethical approval was not required under UK research governance guidelines. The study was conducted in accordance with the principles of the Declaration of Helsinki.

### Provenance and Peer Review

Not commissioned; externally peer reviewed.

### Data Availability Statement

All data used in this study are publicly available. Referral to Treatment data are published by NHS England at https://www.england.nhs.uk/statistics/statistical-work-areas/rtt-waiting-times/. Patient Reported Outcome Measures data are published by NHS England at https://digital.nhs.uk/data-and-information/publications/statistical/patient-reported-outcome-measures-proms. Appointments in General Practice data are published by NHS England at https://digital.nhs.uk/data-and-information/publications/statistical/appointments-in-general-practice. General Practice Patient Survey data are published at https://gp-patient.co.uk.

The CCG-to-ICB linkage was built from the Office for National Statistics Postcode Directory and the NHS England Sub-ICB data dictionary, both publicly available. The provider-to-ICB assignment was built from the Referral to Treatment publication cited above.

### AI Use

During the preparation of this work, the authors used generative AI (ChatGPT) to check for grammar, punctuation errors, and language refinement. After using this tool, the authors reviewed and edited the text as needed and take full responsibility for the content of the publication.

## Appendix

## Appendix A. Ordered Logit Models for Patient Dissatisfaction

**Table A1.**
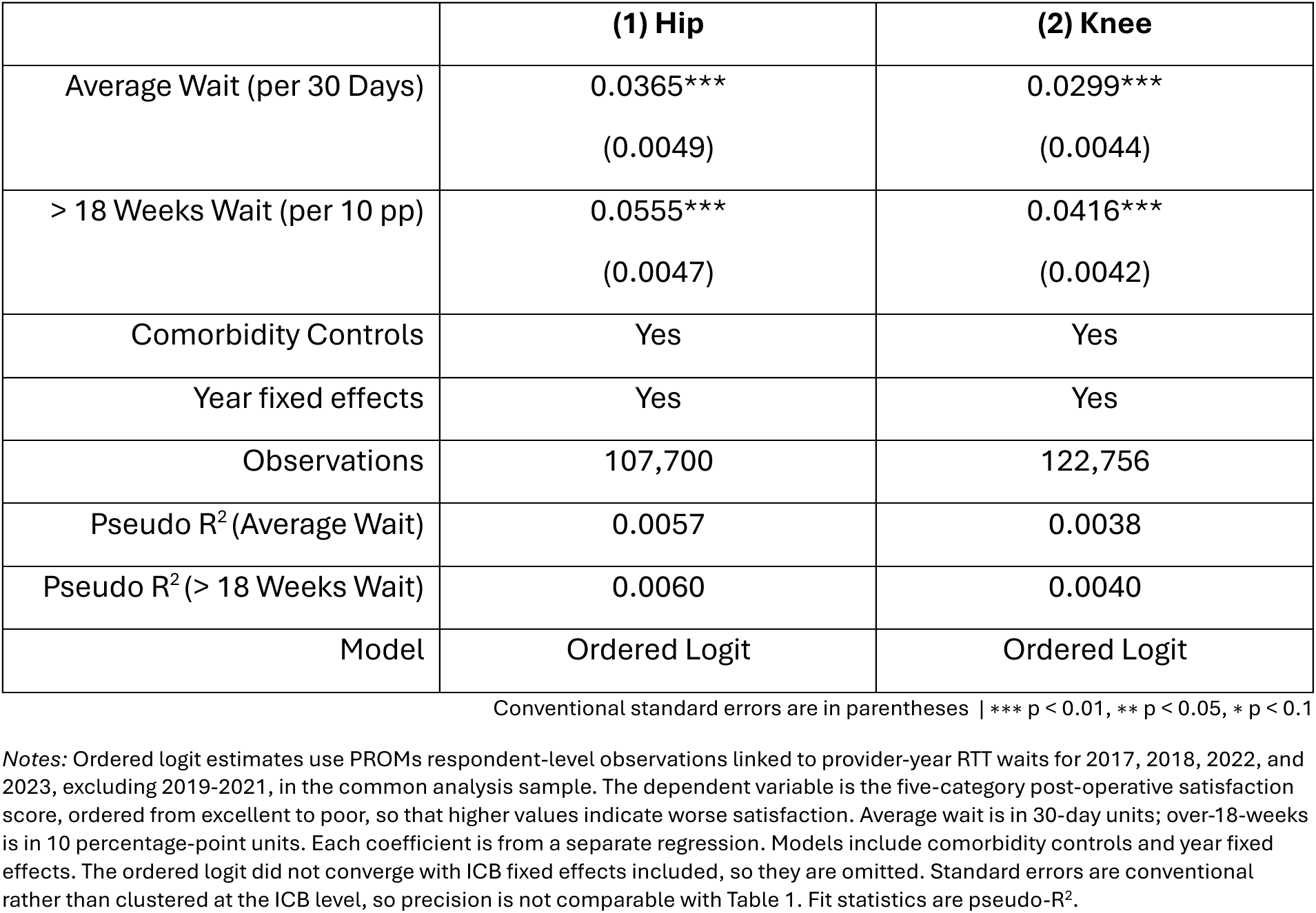
Ordered Logit Regression Estimates of the Association between Waiting Times and Post-Operative Patient Satisfaction Score after Hip and Knee Replacement.

|  | (1) Hip | (2) Knee |
| --- | --- | --- |
| Average Wait (per 30 Days) | 0.0365***<br>(0.0049) | 0.0299***<br>(0.0044) |
| > 18 Weeks Wait (per 10 pp) | 0.0555***<br>(0.0047) | 0.0416***<br>(0.0042) |
| Comorbidity Controls | Yes | Yes |
| Year fixed effects | Yes | Yes |
| Observations | 107,700 | 122,756 |
| Pseudo R<sup>2</sup> (Average Wait) | 0.0057 | 0.0038 |
| Pseudo R<sup>2</sup> (> 18 Weeks Wait) | 0.0060 | 0.0040 |
| Model | Ordered Logit | Ordered Logit |

Conventional standard errors are in parentheses | \*\*\* p < 0.01, \*\* p < 0.05, \* p < 0.1
Notes: Ordered logit estimates use PROMs respondent-level observations linked to provider-year RTT waits for 2017, 2018, 2022, and 2023, excluding 2019-2021, in the common analysis sample. The dependent variable is the five-category post-operative satisfaction score, ordered from excellent to poor, so that higher values indicate worse satisfaction. Average wait is in 30-day units; over-18-weeks is in 10 percentage-point units. Each coefficient is from a separate regression. Models include comorbidity controls and year fixed effects. The ordered logit did not converge with ICB fixed effects included, so they are omitted. Standard errors are conventional rather than clustered at the ICB level, so precision is not comparable with Table 1. Fit statistics are pseudo-R<sup>2</sup>.

## Appendix B. Supplementary ICB-Year Comparison with General Practice

The main analysis uses respondent-level PROMs observations linked to provider-year waits. General practice has no equivalent respondent-level linkage, so the two settings can only be compared after aggregating hip and knee dissatisfaction to the Integrated Care Board by year level. The sample covers all 42 ICBs in England, each observed in all three settings across 2017, 2018, 2022, and 2023, giving 168 observations per setting. This appendix reports that comparison as a supplementary check on whether the wait-dissatisfaction relationship documented for hip and knee replacement extends to a broader care setting.

Figure B1 shows the general practice relationship and Table B1 reports all three settings under a common specification. At this level of aggregation, the knee association remained positive and precisely estimated, the hip association retained the expected positive sign but lost precision, and the general practice association was close to zero. Two features of the design limit how far this can be pushed. The general practice estimate was imprecise, with a confidence interval wide enough to contain both a null and an association of the magnitude estimated for hip and knee replacement. The general practice wait measure also varied little within ICB once ICB and year fixed effects were absorbed, with the binned means in Figure B1 spanning less than two days of residualised wait in total.

**Figure B1.**
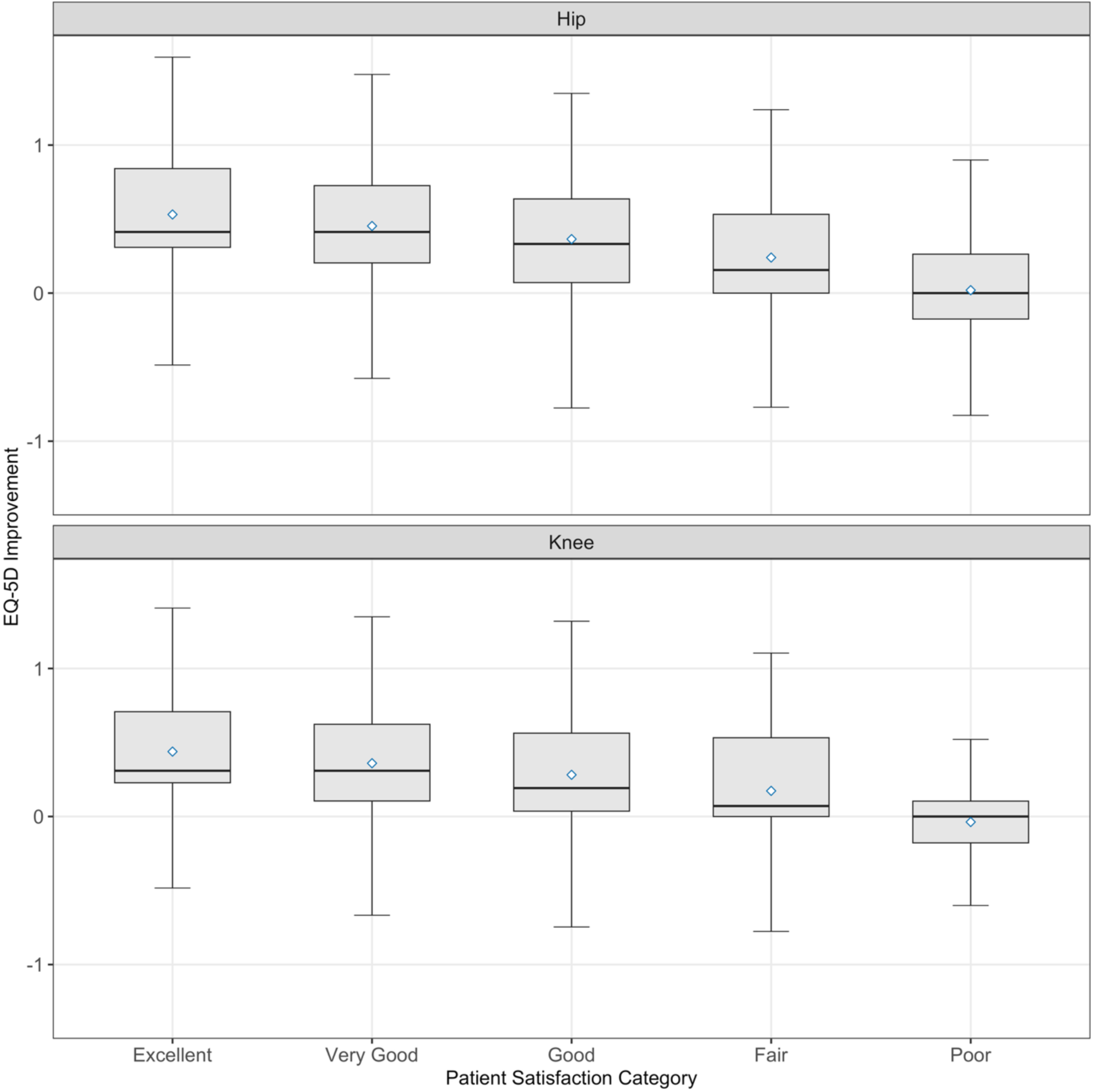
Average Wait and Patient Dissatisfaction for General Practice. *Notes:* Binned scatter plot uses general practice ICB-year observations for 2017, 2018, 2022, and 2023, excluding 2019-2021. Markers are bin means of residualised waiting time and GPPS overall dissatisfaction. Variables are residualised on ICB and year fixed effects. Waits are in 30-day units and dissatisfaction is in percentage points.

**Table B1.** OLS Estimates of the Association between Waiting Times and Patient Dissatisfaction across Hip Replacement, Knee Replacement, and General Practice at the ICB-Year Level.

|  | **(1) Hip** | **(2) Knee** | **(3) General Practice** |
| --- | --- | --- | --- |
| Average Wait (Days) | 0.0048<br>(0.0048) | 0.0131***<br>(0.0048) | -0.0117<br>(0.7170) |
| ICB fixed effects | Yes | Yes | Yes |
| Year fixed effects | Yes | Yes | Yes |
| Weighted | Yes | Yes | Yes |
| Observations | 168 | 168 | 168 |
| R<sup>2</sup> | 0.5700 | 0.7014 | 0.9682 |
| Model | OLS | OLS | OLS |

Standard errors clustered at the ICB level are in parentheses | \*\*\* p < 0.01, \*\* p < 0.05, \* p < 0.1
Notes: OLS estimates use the ICB-year comparison sample for 2017, 2018, 2022, and 2023, excluding 2019-2021, restricted to 42 ICBs observed in hip, knee, and general practice in all included years. Hip and knee outcomes are PROMs dissatisfaction rates from the common analysis sample aggregated to the ICB-year level; the general practice outcome is the GPPS overall dissatisfaction rate at the ICB-year level. The two measures are not defined symmetrically, as PROMs dissatisfaction combines the lowest two of five categories while GPPS dissatisfaction combines the lowest three, so the comparison concerns the association between waits and dissatisfaction rather than the level of dissatisfaction. Regressions are weighted by the number of responses underlying each dissatisfaction rate: PROMs respondents for hip and knee, and GPPS overall-experience responses for general practice. Dissatisfaction is measured in percentage points; average wait is measured in days, so coefficients are per additional day. Figure B1 reports waits in 30-day units, and estimates in this table can be placed on that scale by multiplying by 30. Models include ICB and year fixed effects.

## Appendix C. Waiting Times and EQ-5D Improvement

**Figure C1.**
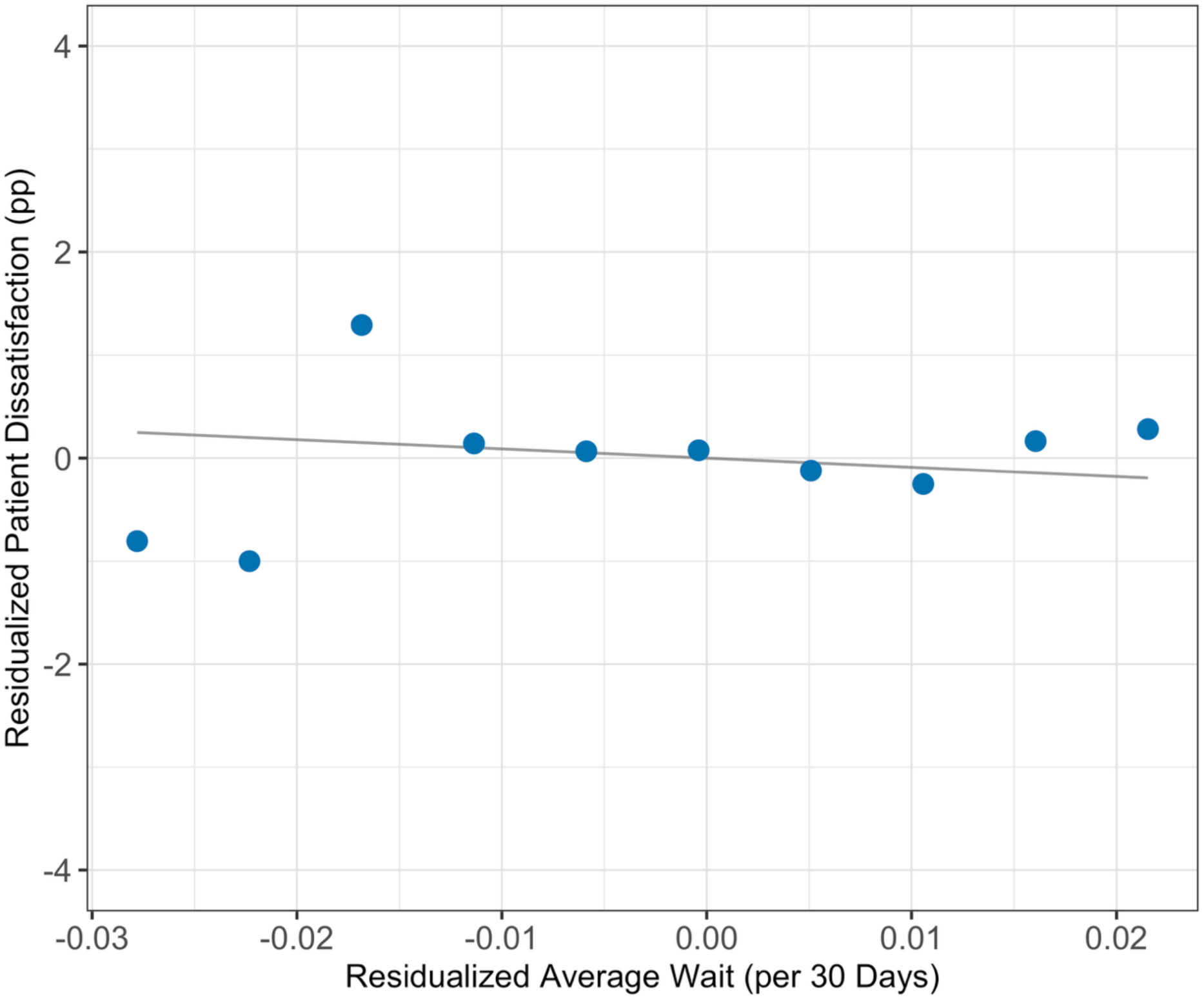
Average Wait and EQ-5D Improvement after Hip and Knee Replacement. *Notes:* Binned scatter plots use PROMs respondent-level observations linked to provider-year RTT waits for 2017, 2018, 2022, and 2023, excluding 2019-2021, comprising the 99,037 hip and 112,792 knee respondents in the common analysis sample with observed EQ-5D improvement. Markers are bin means of residualised waiting time and EQ-5D improvement. Bins with fewer than 20 observations are omitted. Variables are residualised on provider and year fixed effects plus comorbidity controls. EQ-5D improvement is post-operative minus preoperative index. Waits are in 30-day units.

## Appendix D. EQ-5D Improvement by Post-Operative Patient Satisfaction Category

**Figure D1.**
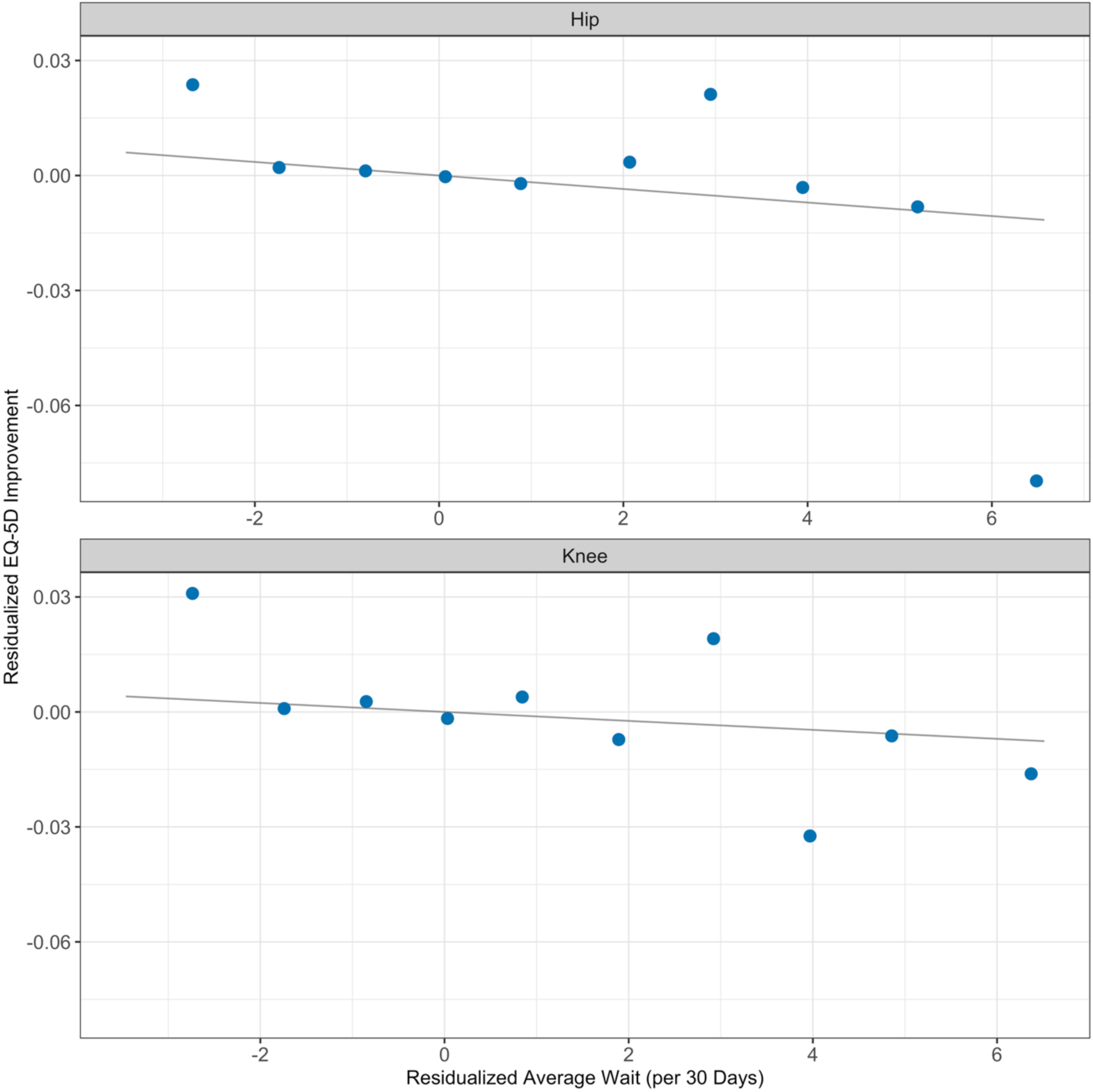
EQ-5D Improvement by Post-Operative Patient Satisfaction Category after Hip and Knee Replacement. *Notes:* Box plots use PROMs respondent-level observations linked to provider-year RTT waits for 2017, 2018, 2022, and 2023, excluding 2019-2021, comprising the 99,037 hip and 112,792 knee respondents in the common analysis sample with observed EQ-5D improvement. EQ-5D improvement is post-operative minus pre-operative index. Boxes show the interquartile range, whiskers show adjacent values, and diamonds show means.

## Appendix E. CCG-to-ICB Mapping and Provider-to-ICB Assignment

Two geographic linkages are required to build the analysis panel. The first maps legacy Clinical Commissioning Group (CCG) identifiers to current Integrated Care Board (ICB) geography and is used to harmonise the Appointments in General Practice and General Practice Patient Survey data across years spanning the NHS commissioning reorganisation. The second assigns each hospital provider a primary ICB and is used for the ICB fixed-effects specification, for the definition of both main hip and knee analysis samples, and for the supplementary comparison against general practice in Appendix B.

### CCG-to-ICB Mapping

The CCG-to-ICB linkage was built from Office for National Statistics Postcode Directory (ONSPD) files. The August 2018 ONSPD file, containing CCG codes, was merged with the May 2025 ONSPD file, containing current Sub-ICB codes, on postcode. The resulting postcode overlaps were linked to ONS CCG and Sub-ICB code and name reference files, then aggregated to CCG-by-Sub-ICB cells.

For each CCG-to-Sub-ICB pair, the number of matched postcodes was counted, and an allocation weight was calculated as the pair’s postcode count divided by the CCG’s total matched postcode count. CCGs that fell entirely within a single current Sub-ICB received a weight of one. CCGs that spanned multiple current Sub-ICBs were retained as multiple weighted rows, rather than being manually assigned to a single destination. ICB and region identifiers were then added from the NHS England Sub-ICB data dictionary, which provides the current Sub-ICB-to-ICB hierarchy.

In the harmonised outputs, legacy CCG-level rows in the Appointments in General Practice and GPPS data are allocated to current ICB geography using these postcode-overlap weights before response counts and appointment counts are summed to the ICB-year level.

### Provider-to-ICB Assignment

Each provider was assigned a single primary ICB using a provider-ICB assignment table constructed from the RTT publication. RTT activity was summed by provider and ICB across the current-ICB era of the publication, which begins in 2020-21, for all ICBs to which the provider was attributed. Each provider was then assigned to the ICB accounting for the largest pooled activity count. Ties were broken first by the number of RTT records and then by ICB code. The assignment was made once over the pooled current-ICB period rather than separately by year, so a provider holds the same ICB in every year in which it appears. Providers were assigned to a single primary ICB rather than allocated proportionally across the ICBs they served.

Providers with no attributable ICB were excluded from the common analysis sample. That exclusion, together with the common outcome and control requirements, accounts for the difference between the linked sample and the sample reported in Tables 1 and 2.

The assignment determines the fixed-effect groups in the dissatisfaction regressions in Table 1, and determines which providers contribute to each ICB-year cell in the supplementary comparison in Appendix B. The Oxford Score specifications in Table 2 used provider fixed effects and did not require ICB attribution to estimate, but the requirement was retained in that sample so that the main hip and knee figures and regression tables rest on aligned sets of providers.

## Appendix F. General Practice Overall Dissatisfaction Harmonisation

The general practice overall dissatisfaction measure used in Figure B1 and Table B1 is constructed from harmonised weighted response counts in the General Practice Patient Survey (GPPS). The GPPS is published annually, with fieldwork undertaken from approximately January to March each year. Survey years are aligned to analysis years as: analysis year equals survey year minus one, so that survey years 2018, 2019, 2023, and 2024 correspond to analysis years 2017, 2018, 2022, and 2023.

Survey results were published at CCG level for retained survey years 2018 and 2019, and at ICB level from survey year 2022 onwards. CCG-level rows were mapped to current ICB geography using the postcode-overlap weights described in Appendix E, and weighted response counts were then summed to the ICB-year level. Retained survey years 2023 and 2024 use the published ICB-level geography directly.

Variables were aligned across survey years using a manually reviewed crosswalk anchored on the 2019 GPPS variable dictionary. The 2019 dictionary was chosen as the base because it is the final retained survey year published at CCG level. The crosswalk maps each year’s source variable codes onto the 2019 base variable and response definitions.

The GPPS variable used is overall experience of the general practice, which asks respondents to rate their experience on a five-point scale from “very good” to “very poor”. The wording, response scale, and dissatisfaction definition are consistent across all survey years in the retained panel. Variable codes changed from Q28_1-Q28_5 in survey years 2018 to 2023 to overallexp_1.count-overallexp_5.count in survey year 2024, but the underlying item is unchanged.

Dissatisfaction is defined as the combined share of respondents answering, “neither good nor poor”, “fairly poor”, or “very poor”. For each ICB-year, weighted dissatisfied responses are summed across these three categories and divided by weighted total responses across all five categories. The analysis uses weighted response counts rather than published percentage variables, and no additional reweighting or imputation is applied. The only weighting introduced by the pipeline is the postcode-overlap allocation weight used to map legacy CCG-level rows to current ICB geography.

